# Impact of accelerating booster vaccination amidst Omicron surge in the United States

**DOI:** 10.1101/2022.01.22.22269655

**Authors:** Pratha Sah, Thomas N Vilches, Seyed M. Moghadas, Meagan C. Fitzpatrick, Eric C Schneider, Alison P. Galvani

**Author notes:** Corresponding author: Alison P. Galvani. Center for Infectious Disease Modeling and Analysis (CIDMA), Yale School of Public Health, New Haven, Connecticut, USA. Authors contributed equally (Pratha Sah, Thomas N Vilches, Seyed M. Moghadas).

## Abstract

COVID-19 infections driven by the Omicron variant are sweeping across the United States. Although early evidence suggests that the Omicron variant may cause less severe disease than previous variants, the explosive spread of infections threatens to drive hospitalizations and deaths to unprecedented high levels, swamping already overburdened hospitals. Booster vaccination appears to be effective at preventing severe illness and hospitalization. However, the pace of booster vaccination in the US has been slow despite the available infrastructure to administer doses at a much higher rate.

We used an age-stratified, multi-variant agent-based model to project the reduction in COVID-related deaths and hospitalizations that could be achieved by accelerating the current daily pace of booster vaccination in the US. We found that doubling the rate of booster vaccination would prevent over 400,000 hospitalizations and 48,000 deaths. Tripling the booster vaccination rate would avert over 600,000 hospitalizations and save 70,000 lives during the first four months of 2022.

## Main text

A confluence of the Delta variant and the emerging Omicron variant is causing over a million new daily cases and rapidly escalating hospitalizations in the United States (US). The Omicron variant was first reported in the US on December 1, 2021, but by the end of 2021 it comprised the majority of new infections and has fueled the largest COVID-19 surge since the beginning of the pandemic.^1^

Early evidence suggests that the Omicron variant may cause less severe disease than infection with previous variants.^2^ However, the high transmissibility and immune-evasive nature of this variant mean that hospitalizations and deaths could nonetheless surpass the peaks observed during the Delta surge in 2021. Fifteen weeks after receiving two doses of the Pfizer-BioNTech vaccine, protection against symptomatic disease caused by Delta and Omicron variants is estimated to be only 72% and 34%, respectively.^3^ Booster dosing appears to raise this protection to 93% and 76%, respectively.^3^

Despite the protective benefits, the pace of booster vaccination in the US has been slow. Among the elderly population 65 years or older, just over 56% of fully vaccinated individuals have received a booster dose. In an effort to improve booster vaccination coverage, the current US administration has introduced several initiatives including Family Mobile Vaccination Clinics through Federal Emergency Management Agency (FEMA) and the addition of over 10,000 vaccination sites across the country, increasing the number of vaccination locations to 90,000.^4^

To evaluate the potential impact of accelerating the booster vaccination rate in the US, we expanded our multi-variant agent-based model of COVID-19 transmission ^5^ by introducing waning immunity and booster vaccination (Appendix). By fitting the model to observed incidence through December 29, 2021, we simulated the pandemic trajectory between January 1 and April 30, 2022. We then compared the projected number of infections, hospitalizations and deaths under continuation of the status quo pace of booster vaccination during December 2021 (approximately 770,000 doses per day) to scenarios that double and triple the daily pace of booster vaccination.

We estimate that between January 1 and April 30, 2022, status quo booster vaccination would lead to approximately 110 million infections, 1,714,329 (95% credible interval [CrI]: 1,597,305 to 1,827,415) hospitalizations, and 218,488 (95% CrI: 195,542 to 244,247) deaths. Doubling the rate of booster vaccination (to ∼ 1.5 million doses per day) would prevent 401,897 (95% CrI: 369,250 to 439,437) hospitalizations and 48,358 (95% CrI: 40,279 to 56,987) deaths. Tripling the booster vaccination rate (to ∼ 2.3 million doses per day) would avert 619,133 (576,113 to 663,140) hospitalizations and save 70,603 (95% CrI: 61,658 to 79,642) lives compared to the status quo (Figure 1A). Increasing the pace of booster vaccination will also substantially reduce the peak number of severe disease outcomes. Specifically, we project that at the current pace of booster vaccination, the daily hospital admissions would peak at approximately 30,000 near the end of January, exceeding 20,000 hospitalizations per day for as much as six weeks. Doubling the pace of booster vaccination will reduce the peak to 25,000 per day. Tripling this rate will further reduce the peak to approximately 21,000 new admissions per day, for a period of less than three weeks. We also evaluated the age-specific impact of booster scenarios. Accelerating booster vaccination is projected to be most impactful in reducing adult hospitalizations, especially among the elderly aged 80 years and older (Figure 1B).

**Figure 1.**
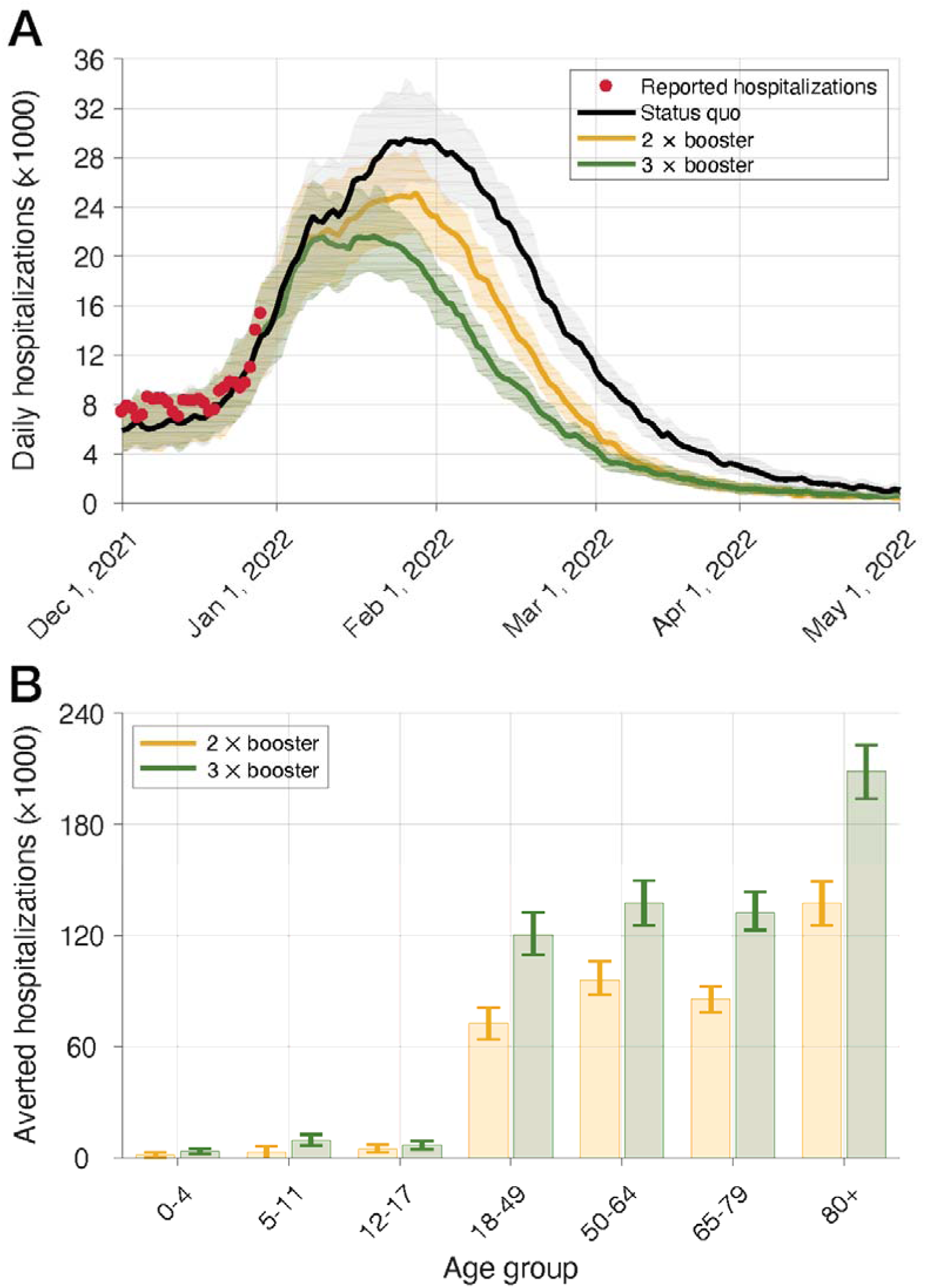
(A) Model projections for daily hospitalizations of the status quo with a daily vaccination rate corresponding to the average pace during December 2021, and accelerated booster programs with double and triple the rate of booster administration from the beginning of January through the end of April 2022. (B) Model projections for the number of hospitalizations averted in different age groups in accelerated booster programs with double and triple the daily rate of booster administration compared to the status quo from the beginning of January through the end of April 2022.

Our results highlight the potential of immediately accelerating booster vaccination to prevent overburdening of hospitals and healthcare workers while curtailing a devastating loss of lives over the next several months. During the initial vaccine rollout in 2021, the average number of vaccine doses administered each topped two million in the US for several weeks. Tripling the rate of booster doses is therefore readily achievable. Booster vaccinations are not only effective in blunting the current surge, but they also bolster otherwise waning population immunity and may protect against future surges.

## Data Availability

All data produced in the present work are contained in the manuscript.

https://github.com/thomasvilches/USomicron/

## Acknowledgments

This work was supported by the Commonwealth Fund. SMM also acknowledges support by the Canadian Institutes of Health Research [OV4 – 170643, COVID-19 Rapid Research] and the Natural Sciences and Engineering Research Council of Canada, Emerging Infectious Disease Modelling, MfPH grant.

## Declaration of interests

The authors declare no competing interests

## Notes

### Competing Interest Statement

The authors have declared no competing interest.

