## Supplementary Information Text for "Impact of accelerating booster vaccination amidst Omicron surge in the United States"

*Authors contributed equally

This Appendix provides details of the model structure and parameterizations for the results reported.

***Model structure***

We employed our previous agent-based model of COVID-19 transmission^1^ and expanded its dynamic structure to account for waning of naturally-acquired or vaccine-elicited immunity, as well as administration of booster vaccination. For fitting the model from the beginning of October 2020 through December 29, 2021, we included the characteristics of four variants in the model, each with a cumulative prevalence of at least 3% in the United States,^2^ including B.1.526 (Iota), B.1.1.7 (Alpha), B.1.617.2 (Delta), and B.1.1.529 (Omicron), in addition to the original Wuhan-I SARS-CoV-2 strain.

The natural history of COVID-19 was modelled with epidemiological classes of individuals as susceptible; latently infected (not yet infectious); asymptomatic (and infectious); pre-symptomatic (and infectious); symptomatic (and infectious) with either mild or severe illness; recovered; and dead. The population was stratified into seven age groups of 0 to 4, 5 to 11, 12-17, 18 to 49, 50 to 64, 65 to 79, and 80+ years based on the US demographics,^3^ and incorporated age-specific risk of hospitalizations and deaths, contact patterns. Daily contacts between individuals were sampled from age-specific negative-binomial distributions with parameters that accounted for the effect of interventions such as isolation of symptomatic cases (Table A1).

**Table A1.** Mixing patterns and the daily number of contacts derived from empirical observations. ^4,5^ Daily numbers of contacts were sampled from negative binomial distributions for different scenarios.

| Age group | Proportion of contacts between age groups | | | | | No. of  daily contacts  Mean (SD) | No. of  daily contacts for isolated individuals  Mean (SD) |
| --- | --- | --- | --- | --- | --- | --- | --- |
|  | 0-4 | 5-19 | 20-49 | 50-65 | 65+ |  |  |
| 0-4 | 0.2287 | 0.1839 | 0.4219 | 0.1116 | 0.0539 | 10.21 (7.65) | 2.86 (2.14) |
| 5-19 | 0.0276 | 0.5964 | 0.2878 | 0.0591 | 0.0291 | 16.793 (11.7201) | 4.70 (3.28) |
| 20-49 | 0.0376 | 0.1454 | 0.6253 | 0.1423 | 0.0494 | 13.795 (10.5045) | 3.86 (2.95) |
| 50-65 | 0.0242 | 0.1094 | 0.4867 | 0.2723 | 0.1074 | 11.2669 (9.5935) | 3.15 (2.66) |
| 65+ | 0.0207 | 0.1083 | 0.4071 | 0.2193 | 0.2446 | 8.0027 (6.9638) | 2.24 (1.95) |

***Transmissibility***

We used reported incidence of COVID-19 cases per 100,000 population in the US to calibrate the model and determine the per-contact transmission probability of the original strain during the pre-symptomatic stage of the disease. The calibration started on October 1, 2020 with a pre-existing immunity against COVID-19 that was included in the model using a probability distribution function, based on the reported incidence from the beginning of pandemic to the end of September 2020 to account for waning immunity over time (Figure A2). At the start of calibration, we assumed that contacts between individuals did not exceed 50% of the pre-pandemic level.^6^ The transmissibilities during asymptomatic, mild symptomatic, and severe symptomatic stages relative to pre-symptomatic stage were set to 26%, 44%, and 89%.^7–9^ The per-contact transmission probability of the Iota and Alpha variants were assumed to be 35% and 50% higher than the original strain. This probability for the Delta variant was assumed to be 30% higher than the Alpha variant.^10^ We considered a 35% higher transmissibility of Omicron compared to Delta based on initial estimates derived from South Africa.^11^

***Distribution of disease stages***

The incubation period for each infected individual was sampled from a log-normal distribution with a mean of 5.2 days.^12^ A proportion of infected individuals progressed to a pre-symptomatic stage^13^ with an infectious period which was sampled from a Gamma distribution with a mean of 2.3 days.^8,14^ The symptomatic disease following the pre-symptomatic stage had an average infectious period of 3.2 days, which was also sampled from a Gamma distribution.^15,16^ The infectious period of individuals who remained asymptomatic was sampled from a Gamma distribution with a mean of 5 days.^15,16^

***Disease outcomes***

We assumed that asymptomatic and mild symptomatic cases recover without hospitalization. Self-isolation was implemented to start for symptomatic cases within 24 hours of symptom onset, reducing their number of daily contacts by an average of 74% (Table A1). Severely ill cases due to primary infection were hospitalized within 2-5 days of symptom onset, ^17,18^ and therefore effectively excluded from the chain of disease transmission. The model was parameterized with rates of intensive care unit (ICU) and non-ICU admissions (Table A2).^19–22^ The risk of hospitalization with the Delta variant was assumed to be 2.26 times higher than that due to infection with Alpha.^22^ We considered a 70% reduction of severe disease due to infection by Omicron compared to Delta.^23,24^ The risk of ICU admission was reduced by 36% in severe cases of Omicron compared to Delta.^24^

**Table A2.** Model parameters associated with hospitalization of severe cases.^19–22^

| Proportion of severe cases hospitalized with one or more comorbidities | | 100% |
| --- | --- | --- |
|  | Non-ICU | 60.4% |
|  | ICU | 39.6% |
| Proportion of severe cases hospitalized without any comorbidities | | 10.8% |
|  | Non-ICU | 75% |
|  | ICU | 25% |

***Vaccination and immune dynamics***

We implemented Vaccination as a two-dose strategy following the recommended schedule of 21 and 28 days between-dose intervals for Pfizer-BioNTech and Moderna vaccines.^25,26^ Combined, these two vaccines constitute ~97% of doses administered in the United States.^27^ The corresponding proportion of these vaccines were 60% and 40% for Pfizer-BioNTech and Moderna vaccines, respectively, with a starting date on December 12, 2020 and a sequential prioritization of (i) healthcare workers (5% of the total population),^28^ adults with comorbidities, and those aged 65 and older; and (ii) other individuals aged 16-64.^29,30^ The number of vaccine doses and distribution of first and the second doses in different age groups were parameterized with the data retrieved from the Vaccine Tracking database of the Center for Disease Control and Prevention (Figure A1).^31^ The minimum age-eligibility for vaccination was 16 and 18 years (for Pfizer-BioNTech and Moderna vaccines, respectively) before May 13, 2021 after which children aged 12 to 15 years became eligible for vaccination. Vaccination of children aged 5-11 years with Pfizer-BioNTech started on November 2, 2021.

We also implemented the booster vaccination with the daily distribution rates reported from August 13, 2021.^31^ The booster eligibility was set to a 6-month period elapsed since the second dose of vaccine. On January 3, 2022, this timeline was reduced to 5 months.^32^

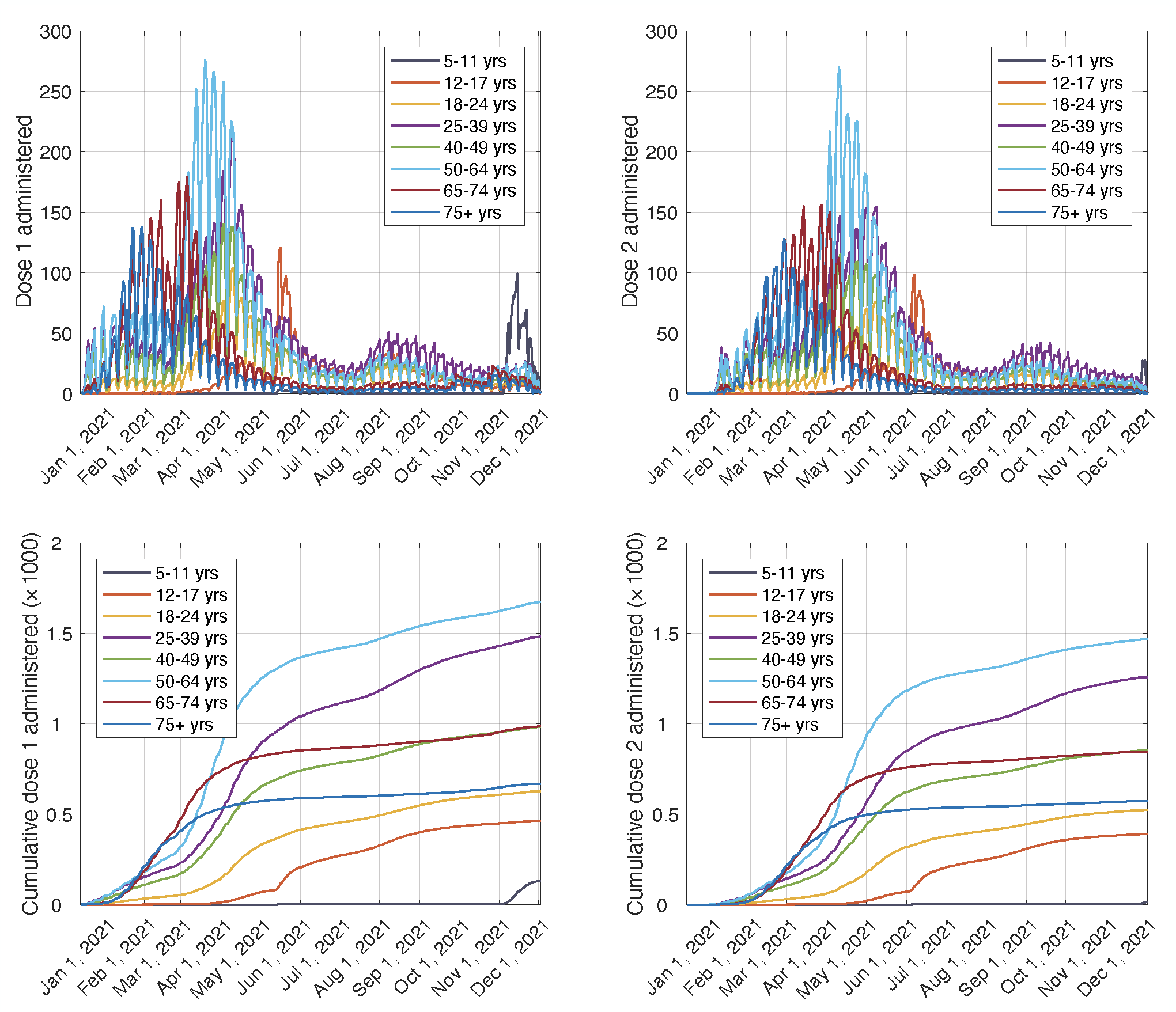

**Figure A1**. Age-specific distribution of vaccine doses per 100,000 population.

**Table A3.** Estimated vaccine efficacies (%) and their 95% confidence intervals from published

Studies for Pfizer-BioNTech vaccines.

| **Vaccine efficacy (%)** | **Timelines** | | **Reference** |
| --- | --- | --- | --- |
| Original strain and Iota variant | 2 weeks after the first dose | 1 week after the second dose | ^26,33–37^ |
| Infection | 46 (40, 51) | 86.1 (82.4, 89.1) |  |
| Symptomatic disease | 57 (50, 63) | 94 (87, 98) |  |
| Severe disease | 62 (39, 80) | 92 (75, 100) |  |
| Alpha variant |  |  | ^38–40^ |
| Infection | 29.5 (22.9, 35.5) | 89.5 (85.9, 92.3) |  |
| Symptomatic disease | 53.6 (50, 63) | 93.7 (91.6, 95.3) |  |
| Severe  disease | 54.1 (26.1, 71.9) | 94 (87, 98) |  |
| Delta variant |  |  | ^40–43^ |
| Infection | 36.8 (32.0, 40.8) | 64 (57, 70) |  |
| Symptomatic disease | 33.5 (20.6, 44.3) | 88 (85.3, 90.1) |  |
| Severe disease | 34 (0, 50) | 80 (73, 85) |  |

**Table A4.** Estimated vaccine efficacies (%) and their 95% confidence intervals from published

Studies for Moderna vaccines.

| **Vaccine efficacy (%)** | **Timelines** | | **Reference** |
| --- | --- | --- | --- |
| Original strain and Iota variant | 2 weeks after the first dose | 1 week after the second dose | ^35,44,45^ |
| Infection | 61 (31, 79) | 93.3  (85.7 - 97.4) |  |
| Symptomatic disease | 92.1  (68.8 - 99.1) | 94.1  (89.3 - 96.8) |  |
| Severe disease | 92.1  (68.8-99.1) | 100 |  |
| Alpha variant |  |  | ^46–48^ |
| Infection | 54.7  (44.8 - 62.9) | 86  (81 - 90.6) |  |
| Symptomatic disease | 88.1  (83.7 - 91.5) | 91  (84 - 95) |  |
| Severe disease | 81.6  (71.0 - 88.8) | 95.7  (73.4 - 99.9) |  |
| Delta variant |  |  | ^46,47^ |
| Infection | 49* | 76  (58 - 87) |  |
| Symptomatic disease | 68* | 70  (45 - 85) |  |
| Severe disease | 78* | 91.6  (81 - 97) |  |

*Assumed: no 95% CI has been documented.

We performed a literature review to derive the efficacy estimates following each dose of vaccine against infection, symptomatic disease, and severe disease for all variants in the model (Tables A3, A4). We assumed that vaccine effectiveness against infection with the immune-evading Omicron variant is reduced by 80%,^49–51^ and booster vaccination mitigates this reduction by 75%.^51^ We also assumed the same degree of immune escape from naturally-acquired infection 12 weeks after recovery from primary infection.^49^ However, natural immunity was associated with 6.7 times lower risk of hospitalization in severe disease compared to fully vaccinated individuals.^52^

To implement the waning immunity after vaccination, we fitted a Gaussian model to estimates of vaccine effectiveness over time, ^48,53,54^ and determined the temporal relative effectiveness curves (Figure A2). The relative effectiveness was used as a multiplicative factor in the efficacy of vaccines after the second dose to determine the temporal immunity of individuals against infection and severe disease for each variant. We applied the same relative effectiveness for waning of naturally-acquired immunity.

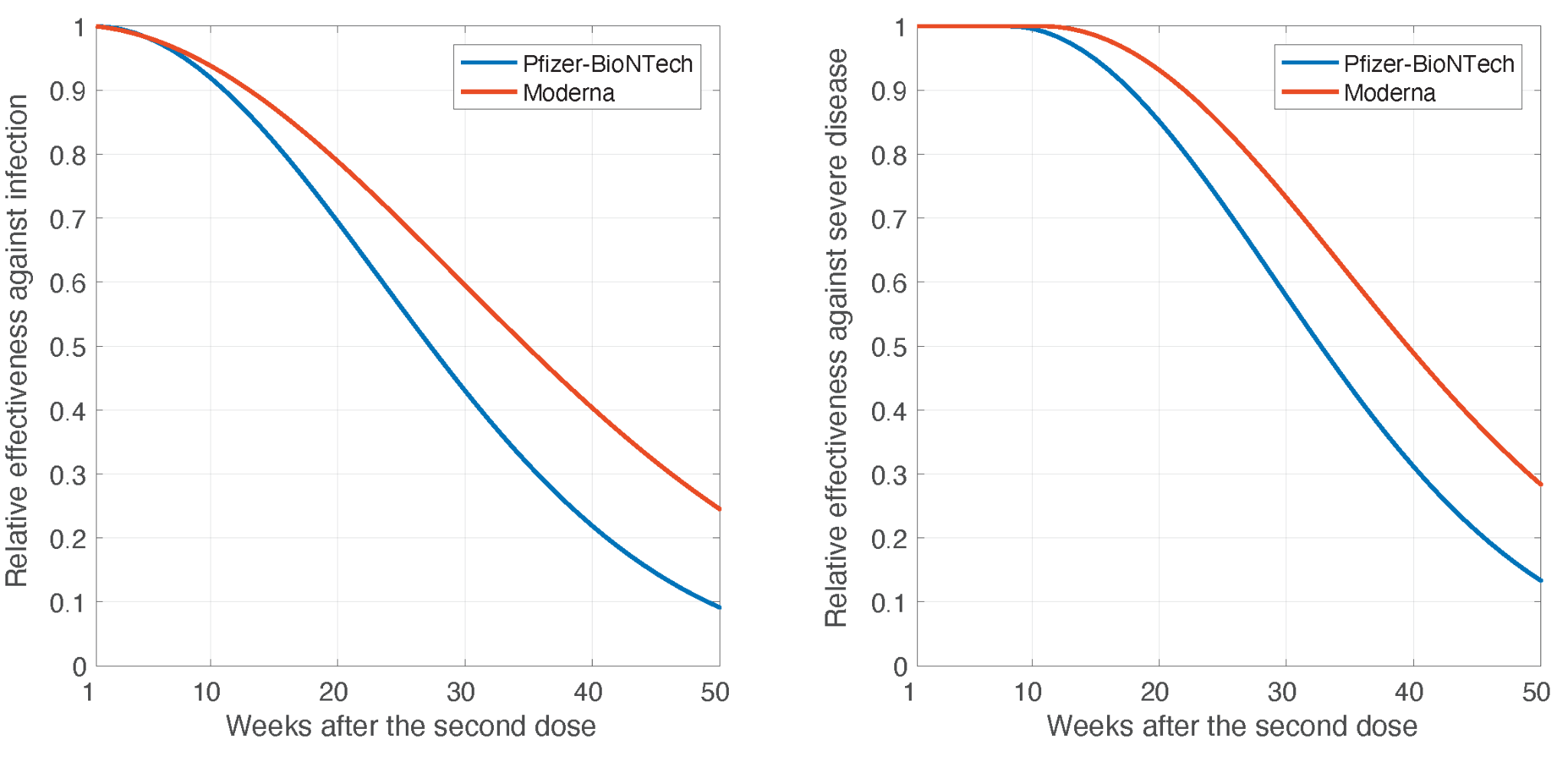

**Figure A2.** Mean of temporal relative effectiveness of vaccines against infection and severe disease derived from a Gaussian fit to estimated effectiveness after the second dose of vaccines ^48,53,54^.

***Model implementation and scenarios***

With the transmission probability derived from the calibration process, we fitted the model to incidence per 100,000 population from October 1, 2020 to November 30, 2021. For fitting, the age-specific contact rates were adjusted throughout the simulations to minimize the difference between the temporal cumulative incidence predicted by the model and the cumulative reported cases, implicitly accounting for the change and effect of various non-pharmaceutical measures. Simulations were averaged over 500 independent Monte-Carlo realizations, and 95% credible intervals derived using a bias-corrected and accelerated bootstrap method (with 500 replications). The model was implemented in Julia, and simulation codes are available at:

[https://github.com/thomasvilches/USomicron/](https://github.com/thomasvilches/USomicron/tree/national_estimation)

To evaluate the impact of an accelerated booster program, we simulated two scenarios in which the daily rate of booster doses was doubled or tripled compared to the average of daily booster doses administered during December 2021 (as status quo). We compared the outcomes of infection, hospitalizations, and deaths under each scenario of the accelerated booster program with the status quo. The projected number of infections, hospitalizations, and death averted in accelerated booster programs are summarized in Table A5. For the scenario of status quo with parameters used in our model, we projected a total of 109,809,406 (95% credible interval [CrI]: 102,533,419 ー 116,984,798) infections from the beginning of January through the end of April 2022, with a peak incidence exceeding 2 million between January 20 and February 5 2022 (Figure A3, A). The projected total hospitalizations and deaths during the same period are 1,714,329 (95% CrI: 1,597,305 ー 1,827,415) and 218,488 (95% CrI: 195,542 ー 244,247), respectively.

**Table A5.** Estimates of COVID-19-attributable infections, hospitalizations, and deaths averted by accelerating COVID-19 booster vaccination program compared to the status quo between January 1, and April 30, 2022.

| **Scenario** | **Doubling daily booster rate** | | **Tripling daily booster rate** | |
| --- | --- | --- | --- | --- |
| **Outcome** | ***Averted*** | ***95% Credible Interval*** | ***Averted*** | ***95% Credible Interval*** |
| Infections | 14,641,734 | 13,436,243 ー 15,759,873 | 23,491,992 | 21,862,199 ー 25,020,954 |
| Hospitalizations | 401,897 | 369,250 ー 439,437 | 619,133 | 576,113 ー 663,140 |
| Deaths | 48,358 | 40,279 ー 56,987 | 70,603 | 61,658 ー 79,642 |

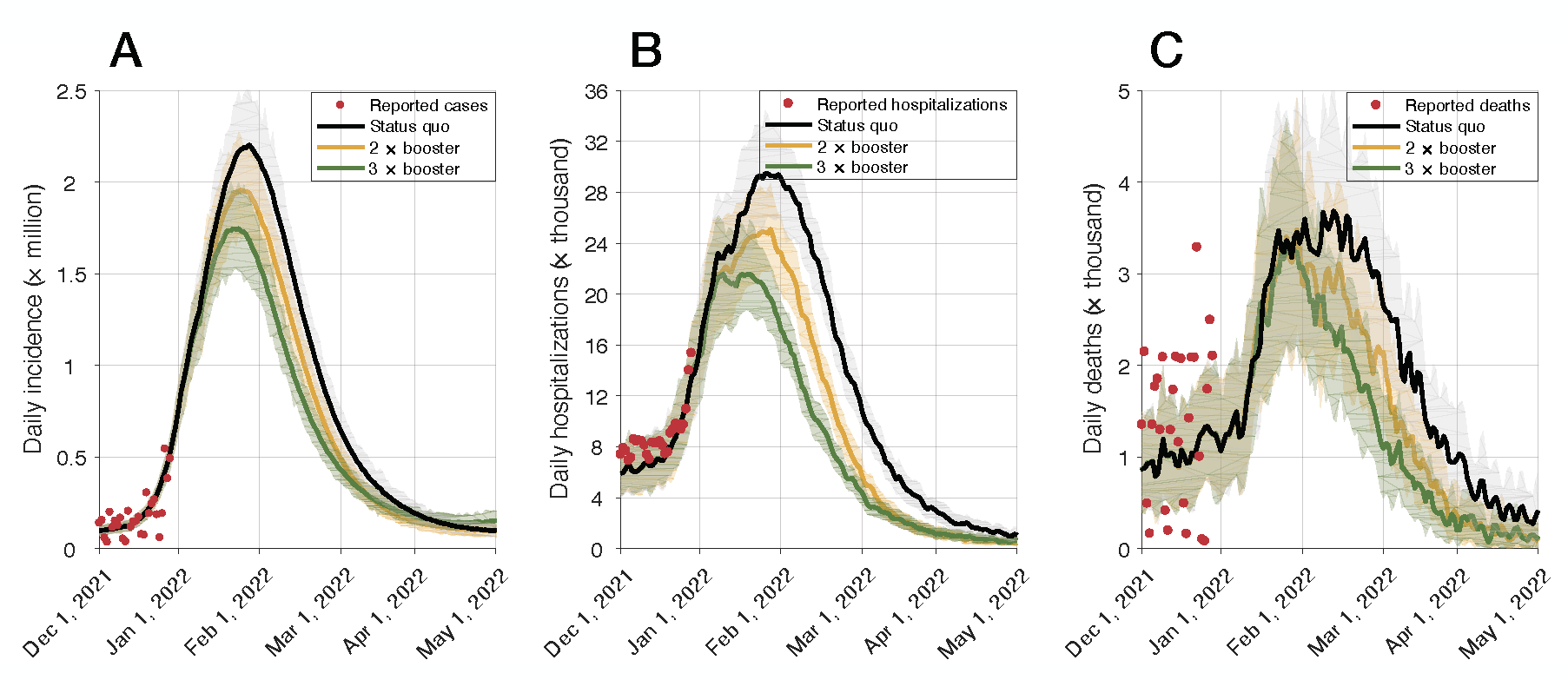

**Figure A3.** Model projections for the incidence of COVID-19 (A), hospitalizations (B), and deaths (C). Simulated scenarios illustrate the projections for status quo with a daily vaccination rate corresponding to average pace during December 2021, and accelerated booster programs with double and triple the rate of booster administration from the beginning of January through the end of April 2022.

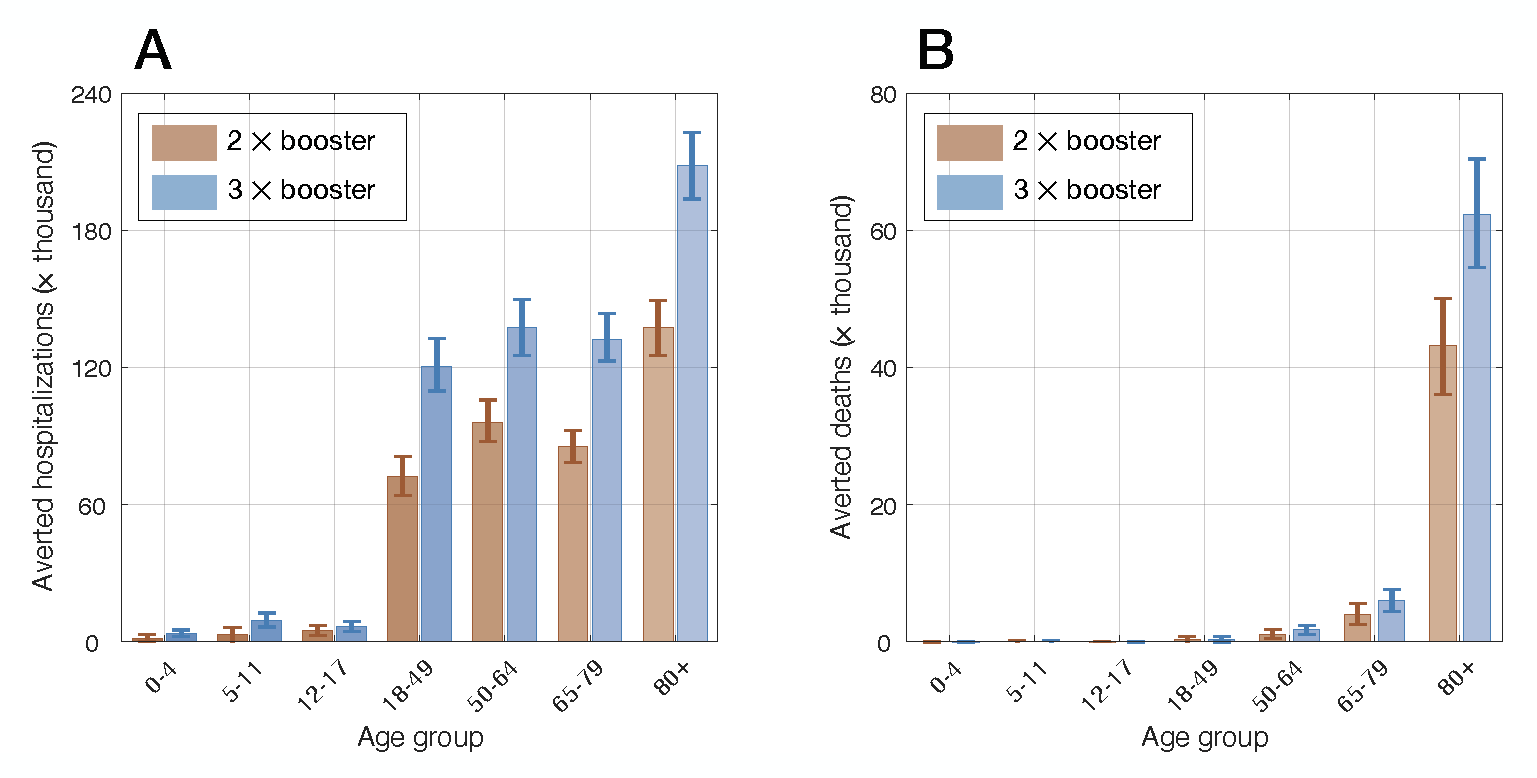

**Figure A4.** Model projections for the number of hospitalizations and deaths averted in different age groups in accelerated booster programs with double and triple the rate of booster administration compared to the status quo from the beginning of January through the end of April 2022.

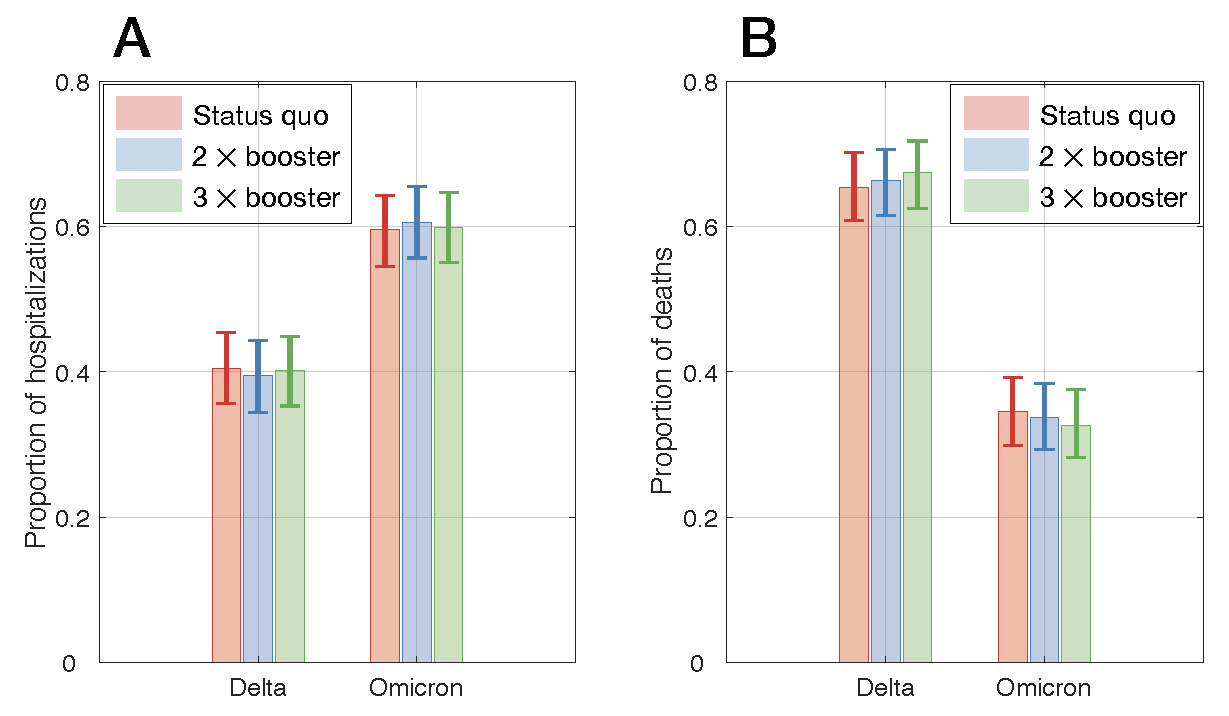

**Figure A5.** Model projections for the proportion of hospitalizations and deaths caused by infection with Delta and Omicron in simulated scenarios of status quo, and accelerated booster programs with double and triple the rate of booster administration from the beginning of January through the end of April 2022.
